# Dim light sensitivity and delayed sleep timing in young people with emerging mental disorders

**DOI:** 10.64898/2026.03.02.26347467

**Authors:** Emiliana Tonini, Ian B. Hickie, Mirim Shin, Joanne S. Carpenter, Alissa Nichles, Natalia Zmicerevska, Elie Jeon, Gabrielle Hindmarsh, Elizabeth Phung, Connie Janiszewski, Tian Lin, Elise M. McGlashan, Sean W. Cain, Jan Scott, Joey W.Y. Chan, Frank Iorfino, Haley M. LaMonica, Yun Ju (Christine) Song, 23andMe Research Team, Naomi R. Wray, Elizabeth M. Scott, Jacob J. Crouse

**Author notes:** Authors contributed equally (Ian B. Hickie, Jacob J. Crouse). **Correspondence:** Dr Emiliana Tonini, Level 5, Moore College, 1 King St, Newtown NSW, 2042, Australia, Brain and Mind Centre, The University of Sydney.

## Abstract

**Background:** Light plays a critical role in mental health, as the primary input to the circadian system, which regulates mood, energy, and the sleep-wake cycle. Altered light sensitivity is a potential mechanism in circadian-associated mental disorders.

**Methods:** Actigraphy-derived sleep, physical activity, and circadian rhythm correlates of the pupillary light reflex were explored in young people with emerging mental disorders. Participants were 27 healthy controls (Mean age=25.67 ± 2.83, 52% female) and 155 young people from the Neurobiology Youth Follow-up Study (Mean age=25.48 ± 5.65; 60% female), recruited from an early intervention mental health service. 32% of the latter group were re-assessed over 12 months. Pupil constriction, average and maximal constriction velocity, and constriction latency were recorded by the PLR-3000 monocular pupillometer in response to dim (~10 lux) and bright (~1500 lux) pulses.

**Results:** Compared to healthy controls, young people with emerging mental disorders had a smaller change in pupil diameter (*p*=0.037) and a slower maximal constriction velocity (*p*=0.018) in response to dim light. In the full sample, decreased dim light sensitivity was correlated with later timing of actigraphy-derived sleep midpoint. Within the clinical cases, increased genetic risk for bipolar disorder was correlated with increased dim light sensitivity, and higher insomnia clinical scores were correlated with decreased dim light sensitivity. Pupillometry measures were stable across time and seasons.

**Conclusion:** Altered light sensitivity may be associated with the emergence of mood disorder in young people and with altered sleep-wake timing.

## 1. Introduction

Light, the primary entraining input of the circadian system (Finger & Kramer, 2021), is recognised as a critical factor in mental health (Crouse et al., 2025; Esaki et al., 2022; Paksarian et al., 2020). While increased exposure to nighttime light has been linked to higher risks of mental disorders and associated symptoms (Burns et al., 2023), evidence suggests that light sensitivity is extremely variable between individuals, with up to 50-fold differences in melatonin suppression to light even in the general population (Phillips et al., 2019). Among young people, who are especially vulnerable to developing mental health conditions (Kessler et al., 2005; McGorry et al., 2024), altered light sensitivity might play a role in the pathophysiology of depressive, anxiety, and bipolar disorders via disturbances in circadian rhythms (Chellappa, 2020; Hickie & Crouse, 2024; Roguski et al., 2024). Quantifying light sensitivity and its clinical and sleep-wake/circadian correlates could offer novel insights into the early detection and differentiation of mood disorder subtypes potentially caused by circadian disturbance (Carpenter et al., 2021; Geoffroy & Maruani, 2025).

Traditionally used to assess parasympathetic and/or sympathetic modulations and providing a metric of the integrity and function of the autonomic nervous system (Hall & Chilcott, 2018), the pupillary light reflex (PLR), a measure of pupil constriction and dilation in response to light, offers one avenue for such investigations. Intrinsically photosensitive retinal ganglion cells (ipRGCs) which transmit light signals to the olivary pretectal nucleus (OPN)—the PLR regulatory centre (Clarke et al., 2003)—also sends a signal to the suprachiasmatic nucleus (SCN), the regulatory centre of the mammalian circadian system (Markwell et al., 2010). Thus, investigating pupillary responses to light allows for the indirect inference of ipRGC “light sensitivity”.

As a relatively quick technique to perform, providing objective physiological measurements, pupillometry is already used to monitor conditions known to affect pupillary dynamics, including neurodegenerative disorders and traumatic brain injuries (Adoni & McNett, 2007; Sparks et al., 2023). Clinical research has increasingly examined pupillometry as a non-invasive diagnostic tool for a range of conditions, including circadian rhythm sleep-wake disorders (McGlashan et al., 2018) and mood disorders (Hall & Chilcott, 2018). However, findings in mood disorders remain mixed and relatively underexplored, particularly in young people. Altered static and dynamic pupil responses in depression, including faster maximal constriction velocity (McCall et al., 2021), slower average and maximal constriction velocity (Mestanikova et al., 2017), and larger dark-adapted pupil diameter (Wang et al., 2014) have been reported relative to healthy controls. In individuals with bipolar disorder, slower pupil dilatation following light exposure has been observed (Biçer et al., 2023). Overall, pupil dynamics appear to differ across populations in which sleep and circadian disturbances are common; however, the direction and clinical relevance of these effects remain unclear (Biçer et al., 2023).

To date, the PLR has been primarily investigated in relation to central autonomic activity and emotion regulation. But little is known about the associations between actigraphy-derived objective sleep-wake and circadian parameters and the PLR, as proxy measure of circadian light sensitivity, in the context of youth mental health, as well as the status of the PLR as a state vs trait measure. Thus, this study aims (1) to compare PLR measures between young people with emerging mental disorders and healthy controls (recruited from the same research centre), (2) to investigate the correlations between the PLR and objective sleep-wake and circadian parameters measured by actigraphy, clinical scales, and polygenic risk scores for major mental disorders and sleep/circadian traits, and (3) to investigate the longitudinal stability and seasonal patterns of PLR measures. By identifying these relationships, we aim to advance our understanding of the role of light sensitivity in mood disorders and to examine whether pupillometry might be a useful clinical tool for early detection and differential diagnosis of mental disorder subtypes.

## 2. Methods

### 2.1. Participants

Participants were 155 young people from the *Neurobiology Youth Follow-up Study* (Nichles et al., 2021) (Mean age=25.5, SD=5.7; 60% Female), who were seeking help for mental health-related problems at an early intervention service (*headspace)* in Sydney, Australia. The study is a prospective clinical cohort study with repeated measures at baseline and 6-month, 1-year, 2-year, and 3-year follow-ups. Self-report questionnaires, clinician-rated scales, a neuropsychological battery, pupillometry, and biological measures were collected (e.g. QIDS, PSQI, GAD-7). Recruitment was not limited by specific diagnoses. Young people who were not proficient in English language or who had a clinically assessed intellectual disability were ineligible due to the difficulty to complete the assessments and/or to provide informed consent. The study commenced in 2021 and the pupillometry protocol was introduced in 2024. As of December 2025, forty-nine participants had pupillometry data at two time points, and 11 at three time points.

Healthy controls (HCs) were 27 young adults (Mean age=25.67, SD=2.83; 52% female) from a related study examining circadian biomarkers and consented to their data being used in any or related research studies. HCs were excluded if they self-reported a diagnosis of or treatment for a mental health condition and/or their self-ratings indicated a score above the established thresholds on validated screening measures of depressive (PHQ-9), anxiety (GAD-7), or psychotic-like (PQ-16) symptoms; or if they exhibited an irregular sleep-wake cycle evidence by actigraphy or self-report (PSQI). The clinical and HC samples underwent an identical pupillometry and actigraphy protocol. Details on light exposure calculation and symptom scales are reported in the Supplementary Materials.

Ethics approval was obtained from the Sydney Local Health District Human Research Ethics Committee (2020/ETH01272) for the Neurobiology study and (X23-0148 & 2023/ETH006188) for control participants. All participants provided written informed consent; for those under 16□years, parental/guardian consent was also obtained.

### 2.2. Actigraphy

Participants wore a wrist-worn actigraphy recording device (GENEActiv, Activinsights, Kimbolton, UK) for 5-24 days (median=13 days; 75% had 10-15 days of recording) in their usual environments. Participants were instructed to wear the device on their non-dominant wrist and to only remove it for showering, bathing, or swimming. Data were collected at 30Hz. Raw GENEActiv actigraphy data were processed using an open-source R package, GGIR (version 3.0.5), developed to process multiday accelerometer data (Migueles et al., 2019). Post-processing was completed using the package mMARCH.AC (version 2.9.2) according to protocols established for the ‘Mobile Motor Activity Research Consortium for Health’ (mMARCH) (Guo et al., 2022). Any days with <8 hours of data (i.e., non-wear) were removed. Missing data on remaining days were imputed with the mean of that epoch on all other days. GGIR and mMARCH.AC algorithms were used to generate sleep (Van Hees et al., 2018), physical activity (Van Hees et al., 2013), and circadian rhythm parameters (Guo et al., 2022). Actigraphy data were available for 101 young people with emerging mental disorders and 24 controls. The main reasons for the lack of actigraphy data included (1) <5 days of valid data; or (2) device malfunction. Descriptive data of the sample with complete actigraphy data are reported in Table S1.

The following actigraphy measures were selected for analysis on the basis of their common use in studies of mood disorders: sleep onset; sleep offset; sleep duration (number of minutes estimated as asleep per 24h period); sleep efficiency (percentage of time estimated as asleep between sleep onset and wake time); sleep midpoint (the halfway point between sleep onset and wake time); relative amplitude (RA); intradaily variability (IV); inter-daily stability (IS); Sleep Regularity Index (SRI); total activity count (TAC); and moderate-to-vigorous physical activity (MVPA).

SRI, a metric comparing the similarity of sleep patterns from one day to the next, was calculated using the R package sleepreg (version 1.3.5) (Windred et al., 2024) based on sleep-wake estimates by epoch from GENEActiv GGIR output. A minimum of 5 overlapping days was required to calculate the SRI score.

### 2.3. Polygenic risk scores

Saliva was collected at the Brain and Mind Centre (BMC) at the University of Sydney or the participants’ home using the Oragene saliva self-collection tube and mailed inside a specimen carrier bag in a pre-addressed padded envelope for processing at the Human Studies Unit (HSU) at the Institute of Molecular Bioscience, University of Queensland using the Illumina GSAMDv3 array.

Genotypes were clustered against a large in-house Illumina GSAMDv3 reference set using GenomeStudio v2.0. Quality control (QC) procedures applied across multiple genotyping projects at the HSU were used to arrive at a high-quality set of variants meeting the following requirements: autosomal region, missingness < 0.05, Hardy-Weinberg P > 10^-6^, and minor allele frequency > 0.05 (after QC, 7 samples and 200,796 variants were excluded, leaving 232 samples and 487,747 variants for downstream processing/analysis). Samples were imputed against the Haplotype Reference Consortium (Haplotype Reference Consortium, 2016) panel using IMPUTE v5.0 and poorly imputed variants with information score <0.3 were excluded. Projection of principal components from the 1000 Genomes (Abecasis et al., 2010) reference set onto the genotyped samples were used to identify and excluded a small number (N=24) of individuals of inferred non-European ancestry. Polygenic score (PGS) scoring weights were derived from publicly available genome-wide association studies using GCTB v2.5.2 SBayesRC method (Zheng et al., 2024) and applied scoring weights to imputed data to calculate PGS for bipolar disorder (Mullins et al., 2021), major depressive disorder (Adams et al., 2025), sleep midpoint (Jones et al., 2019), sleep duration (Jansen et al., 2019), chronotype (Jones et al., 2019), insomnia (Jiang et al., 2021), and vitamin D (Revez et al., 2020). PGS were standardized against the ASPREE cohort (McNeil et al., 2019).

### 2.4. Pupillometry

Participants underwent a 20-min pupillometry assessment to examine the pupillary light reflex (PLR). The PLR was measured by a handheld PLR-3000 Monocular Pupillometer (NeurOptics, California, USA) in a dark, windowless room (<2 lux, verified using a Smart Sensor Digital Lux; model AS803) at the BMC. The pupillometer is a monocular single-camera system that presents light stimuli (from 14 white light-emitting diodes [LEDs]) and tracks responses of the pupil. Participants were adapted to darkness (<2 lux) for 10-minutes, with the screen of the device (viewable only to the researcher) filtered by neutral density filters. The PLR was measured by delivering two 1-second light pulses (dim [~10 lux] followed by bright [~1500 lux]). A measure of baseline pupil size (for calculation of change metrics) was taken for 3-seconds before the onset of light stimuli for each measurement. All measurements were taken on the left eye and in daytime hours between 2-8 hours after habitual wake time (for cases: average assessment time=1:05pm [range=9:00am-5:01pm]; for controls: average assessment timing=12:54pm [range=10:43am-3:57pm]). Participants were instructed not to wear contact lenses or eye makeup.

The following metrics were examined: static pupil diameter, and then in the two light conditions (dim and bright): change of pupil size (amplitude); average constriction velocity (ACV); maximal constriction velocity (MCV), and constriction latency (CL). Measurements are in mm or mm/s and are automatically calculated by the device during successful assessment. A correlation matrix between these PLR metrics is reported in Figure S2.

Values above Q3 + 3*IQR and below Q3 – 3*IQR were classified as extreme outliers and removed (N=55 values across 31 participants [i.e. 2.6% of all measures]). Subsequently, missing PLR values were imputed using the *imputePCA* function from the *missMDA* R package (version 1.19). A Principal Component Analysis (PCA) was then run to obtain a single composite measure that allowed a statistical summary of these PLR metrics. A separate PCA was run for the dim and the bright light condition at each visit. In each model, the number of PCs explaining up to 80% of the variance in the dim condition and in the bright condition respectively, were combined into a single score and referred to hereafter as “Dim light sensitivity” and “Bright light sensitivity”.

### 2.5. Statistical analysis

Analyses were conducted in R (version 4.3.0). Demographic characteristics, self-rating scale scores, actigraphy measures, and pupillometry measures were compared between cases and healthy controls using an F-test or Chi-square, as appropriate. Pupillometry and actigraphy comparisons were adjusted for the effect of age. First, we calculated partial correlation coefficients between pupillometry measures and actigraphy-derived sleep, physical activity, and circadian rhythm measures in the whole sample, accounting for the effect of age and the static pupil diameter.

Then, we explored a series of within-cases analyses. First, we investigated whether (1) associations between pupillometry and actigraphy measures were mediated by objective light exposure measures. Then, we calculated partial correlations (2) between pupillometry measures and clinical scale scores, accounting for the effect of age and static pupil diameter and (3) between pupillometry measures and PGS, accounting for the effect of population ancestry with two PCs, age, and static pupil diameter. Next, we compared (4) pupillometry measures between those who were prescribed vs not-prescribed mood stabilizers (given the potential role of mood stabilizers such as lithium in altering light sensitivity) (Roguski et al., 2024), and (5) those who were prescribed SSRI or SNRI vs not-prescribed either SSRI nor SNRI, using analyses of covariance and accounting for the effect of age. Finally, for cases with longitudinal time points of pupillometry, we investigated (6) the stability of PLR measures across time and then (7) seasonal variation. The longitudinal stability of PLR measures was investigated using a linear mixed-effects model using the *lme4* (version 1.1) and *lmerTest* R packages (version 3.1), with “visit” entered as a fixed effect and a random intercept for participant. Separate models were estimated for the dim and the bright light conditions. Pupillometry measures were compared between seasons using a series of one-way ANOVAs. Due to the exploratory nature of these preliminary analyses, we did not correct for multiple comparisons.

## 3. Results

### 3.1. Comparison of Cases and Healthy Controls

Age and sex distributions were not significantly different between cases and HCs. When comparing pupillometry measures, the change of pupil size after a dim light (~10 lux) pulse was significantly smaller in cases (Amplitude=1.73mm) compared to HCs (Amplitude=1.93mm) (F[1,179]=4.448; *p*=0.037) and the maximal velocity of pupil constriction after a dim light pulse was significantly slower in cases (MCV=4.35mm/s) compared to HCs (MCV=4.75mm/s) (F[1,179]=5.762; *p*=0.018). There were no differences in actigraphy-derived measures between HCs and the cases. Detailed analyses are reported in Table 1 and Figure 1.

**Table 1.**
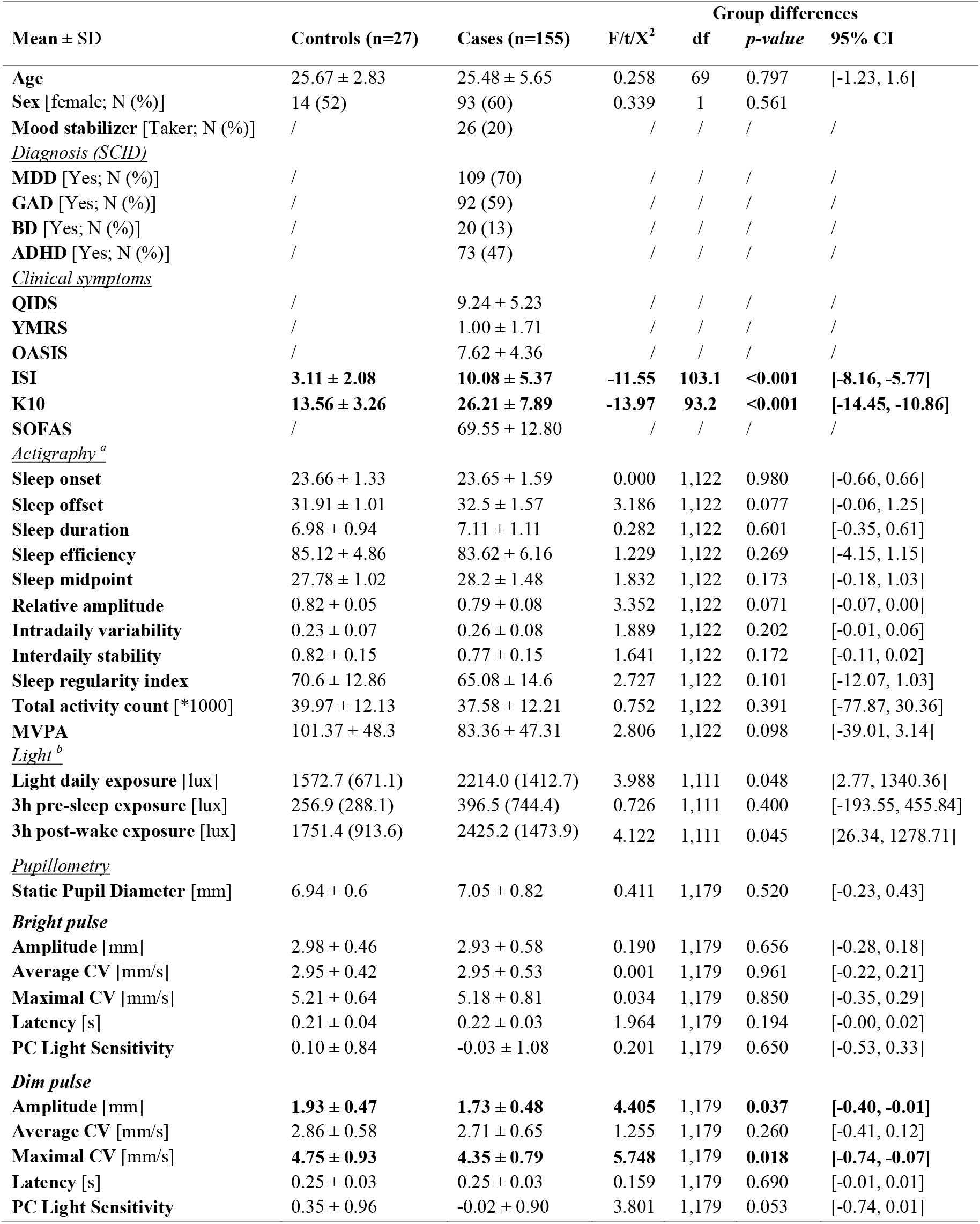
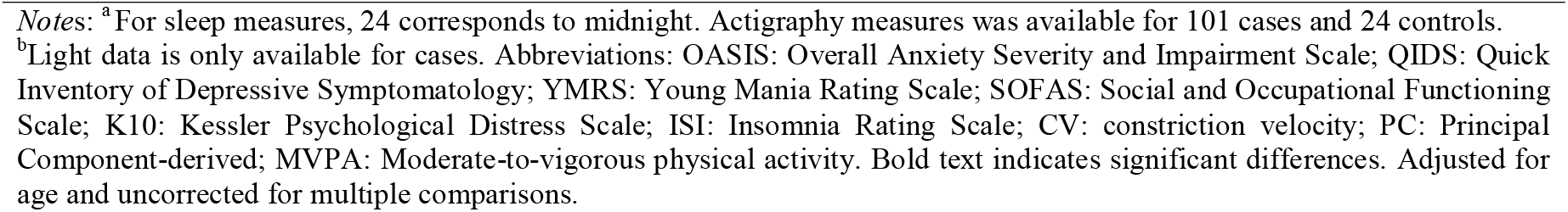
Descriptive statistics of the samples at the first visit.

| Mean ± SD | Controls (n=27) | Cases (n=155) | F/t/X<sup>2</sup> | Group differences | | |
| --- | --- | --- | --- | --- | --- | --- |
|  |  |  |  | df | p-value | 95% CI |
| **Age** | 25.67 ± 2.83 | 25.48 ± 5.65 | 0.258 | 69 | 0.797 | [-1.23, 1.6] |
| **Sex** [female; N (%)] | 14 (52) | 93 (60) | 0.339 | 1 | 0.561 |  |
| **Mood stabilizer** [Taker; N (%)] | / | 26 (20) | / | / | / | / |
| <u>Diagnosis (SCID)</u> |  |  |  |  |  |  |
| **MDD** [Yes; N (%)] | / | 109 (70) | / | / | / | / |
| **GAD** [Yes; N (%)] | / | 92 (59) | / | / | / | / |
| **BD** [Yes; N (%)] | / | 20 (13) | / | / | / | / |
| **ADHD** [Yes; N (%)] | / | 73 (47) | / | / | / | / |
| <u>Clinical symptoms</u> |  |  |  |  |  |  |
| **QIDS** | / | 9.24 ± 5.23 | / | / | / | / |
| **YMRS** | / | 1.00 ± 1.71 | / | / | / | / |
| **OASIS** | / | 7.62 ± 4.36 | / | / | / | / |
| **ISI** | **3.11 ± 2.08** | **10.08 ± 5.37** | **-11.55** | **103.1** | **<0.001** | **[-8.16, -5.77]** |
| **K10** | **13.56 ± 3.26** | **26.21 ± 7.89** | **-13.97** | **93.2** | **<0.001** | **[-14.45, -10.86]** |
| **SOFAS** | / | 69.55 ± 12.80 | / | / | / | / |
| <u>Actigraphy<sup>a</sup></u> |  |  |  |  |  |  |
| **Sleep onset** | 23.66 ± 1.33 | 23.65 ± 1.59 | 0.000 | 1,122 | 0.980 | [-0.66, 0.66] |
| **Sleep offset** | 31.91 ± 1.01 | 32.5 ± 1.57 | 3.186 | 1,122 | 0.077 | [-0.06, 1.25] |
| **Sleep duration** | 6.98 ± 0.94 | 7.11 ± 1.11 | 0.282 | 1,122 | 0.601 | [-0.35, 0.61] |
| **Sleep efficiency** | 85.12 ± 4.86 | 83.62 ± 6.16 | 1.229 | 1,122 | 0.269 | [-4.15, 1.15] |
| **Sleep midpoint** | 27.78 ± 1.02 | 28.2 ± 1.48 | 1.832 | 1,122 | 0.173 | [-0.18, 1.03] |
| **Relative amplitude** | 0.82 ± 0.05 | 0.79 ± 0.08 | 3.352 | 1,122 | 0.071 | [-0.07, 0.00] |
| **Intradaily variability** | 0.23 ± 0.07 | 0.26 ± 0.08 | 1.889 | 1,122 | 0.202 | [-0.01, 0.06] |
| **Interdaily stability** | 0.82 ± 0.15 | 0.77 ± 0.15 | 1.641 | 1,122 | 0.172 | [-0.11, 0.02] |
| **Sleep regularity index** | 70.6 ± 12.86 | 65.08 ± 14.6 | 2.727 | 1,122 | 0.101 | [-12.07, 1.03] |
| **Total activity count** [*1000] | 39.97 ± 12.13 | 37.58 ± 12.21 | 0.752 | 1,122 | 0.391 | [-77.87, 30.36] |
| **MVPA** | 101.37 ± 48.3 | 83.36 ± 47.31 | 2.806 | 1,122 | 0.098 | [-39.01, 3.14] |
| <u>Light<sup>b</sup></u> |  |  |  |  |  |  |
| **Light daily exposure** [lux] | 1572.7 (671.1) | 2214.0 (1412.7) | 3.988 | 1,111 | 0.048 | [2.77, 1340.36] |
| **3h pre-sleep exposure** [lux] | 256.9 (288.1) | 396.5 (744.4) | 0.726 | 1,111 | 0.400 | [-193.55, 455.84] |
| **3h post-wake exposure** [lux] | 1751.4 (913.6) | 2425.2 (1473.9) | 4.122 | 1,111 | 0.045 | [26.34, 1278.71] |
| <u>Pupillometry</u> |  |  |  |  |  |  |
| **Static Pupil Diameter** [mm] | 6.94 ± 0.6 | 7.05 ± 0.82 | 0.411 | 1,179 | 0.520 | [-0.23, 0.43] |
| <u>Bright pulse</u> |  |  |  |  |  |  |
| **Amplitude** [mm] | 2.98 ± 0.46 | 2.93 ± 0.58 | 0.190 | 1,179 | 0.656 | [-0.28, 0.18] |
| **Average CV** [mm/s] | 2.95 ± 0.42 | 2.95 ± 0.53 | 0.001 | 1,179 | 0.961 | [-0.22, 0.21] |
| **Maximal CV** [mm/s] | 5.21 ± 0.64 | 5.18 ± 0.81 | 0.034 | 1,179 | 0.850 | [-0.35, 0.29] |
| **Latency** [s] | 0.21 ± 0.04 | 0.22 ± 0.03 | 1.964 | 1,179 | 0.194 | [-0.00, 0.02] |
| **PC Light Sensitivity** | 0.10 ± 0.84 | -0.03 ± 1.08 | 0.201 | 1,179 | 0.650 | [-0.53, 0.33] |
| <u>Dim pulse</u> |  |  |  |  |  |  |
| **Amplitude** [mm] | **1.93 ± 0.47** | **1.73 ± 0.48** | **4.405** | 1,179 | **0.037** | **[-0.40, -0.01]** |
| **Average CV** [mm/s] | 2.86 ± 0.58 | 2.71 ± 0.65 | 1.255 | 1,179 | 0.260 | [-0.41, 0.12] |
| **Maximal CV** [mm/s] | **4.75 ± 0.93** | **4.35 ± 0.79** | **5.748** | 1,179 | **0.018** | **[-0.74, -0.07]** |
| **Latency** [s] | 0.25 ± 0.03 | 0.25 ± 0.03 | 0.159 | 1,179 | 0.690 | [-0.01, 0.01] |
| **PC Light Sensitivity** | 0.35 ± 0.96 | -0.02 ± 0.90 | 3.801 | 1,179 | 0.053 | [-0.74, 0.01] |

*Notes:* <sup>a</sup> For sleep measures, 24 corresponds to midnight. Actigraphy measures was available for 101 cases and 24 controls.
<sup>b</sup>Light data is only available for cases. Abbreviations: OASIS: Overall Anxiety Severity and Impairment Scale; QIDS: Quick Inventory of Depressive Symptomatology; YMRS: Young Mania Rating Scale; SOFAS: Social and Occupational Functioning Scale; K10: Kessler Psychological Distress Scale; ISI: Insomnia Rating Scale; CV: constriction velocity; PC: Principal Component-derived; MVPA: Moderate-to-vigorous physical activity. Bold text indicates significant differences. Adjusted for age and uncorrected for multiple comparisons.

**Figure 1.**
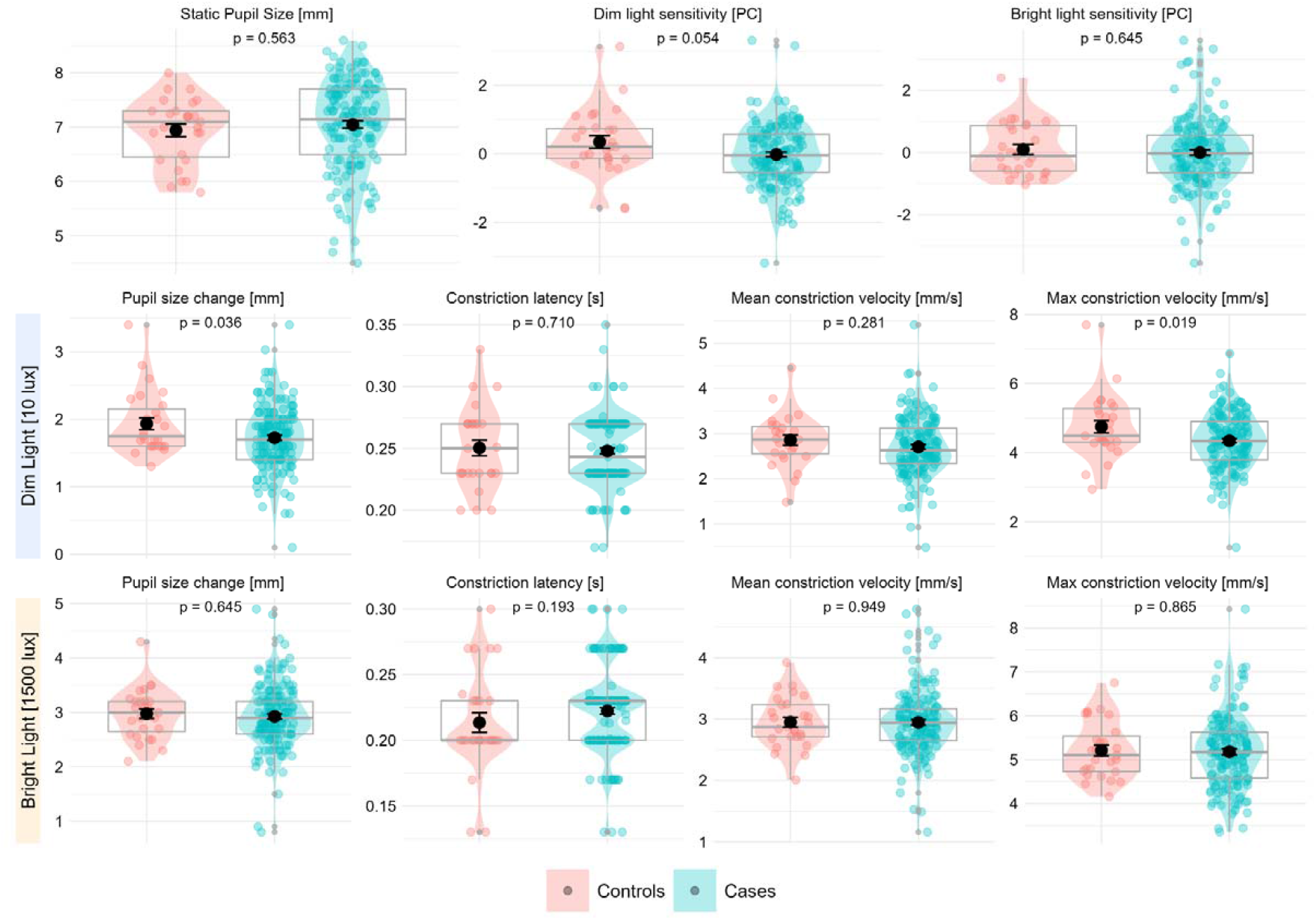
Comparisons of pupillometry measures between clinical cases (N=155) and healthy controls (N=27). Note: In each plot, the box represents the interquartile range with median as line inside the box and mean as a black dot with ± 1 standard error as error bars. Adjusted for effect of age.

### 3.2 Associations between pupillometry and actigraphy measures across all participants

On average, correlations coefficients between actigraphy and pupillometry measures were low-to-moderate. In the whole sample, the increased average constriction velocity after a dim light pulse was correlated with earlier sleep midpoint (r=-0.21; *p*=0.022) and sleep offset (r=-0.21; *p*=0.020), and the increased maximal constriction velocity after a dim light pulse was correlated to earlier sleep offset (r=-0.18; *p*=0.049), indicating that those with slower constriction velocity after a dim light pulse had a later sleep midpoint and later sleep onset. The correlation matrix is presented in Figure 2.

**Figure 2.**
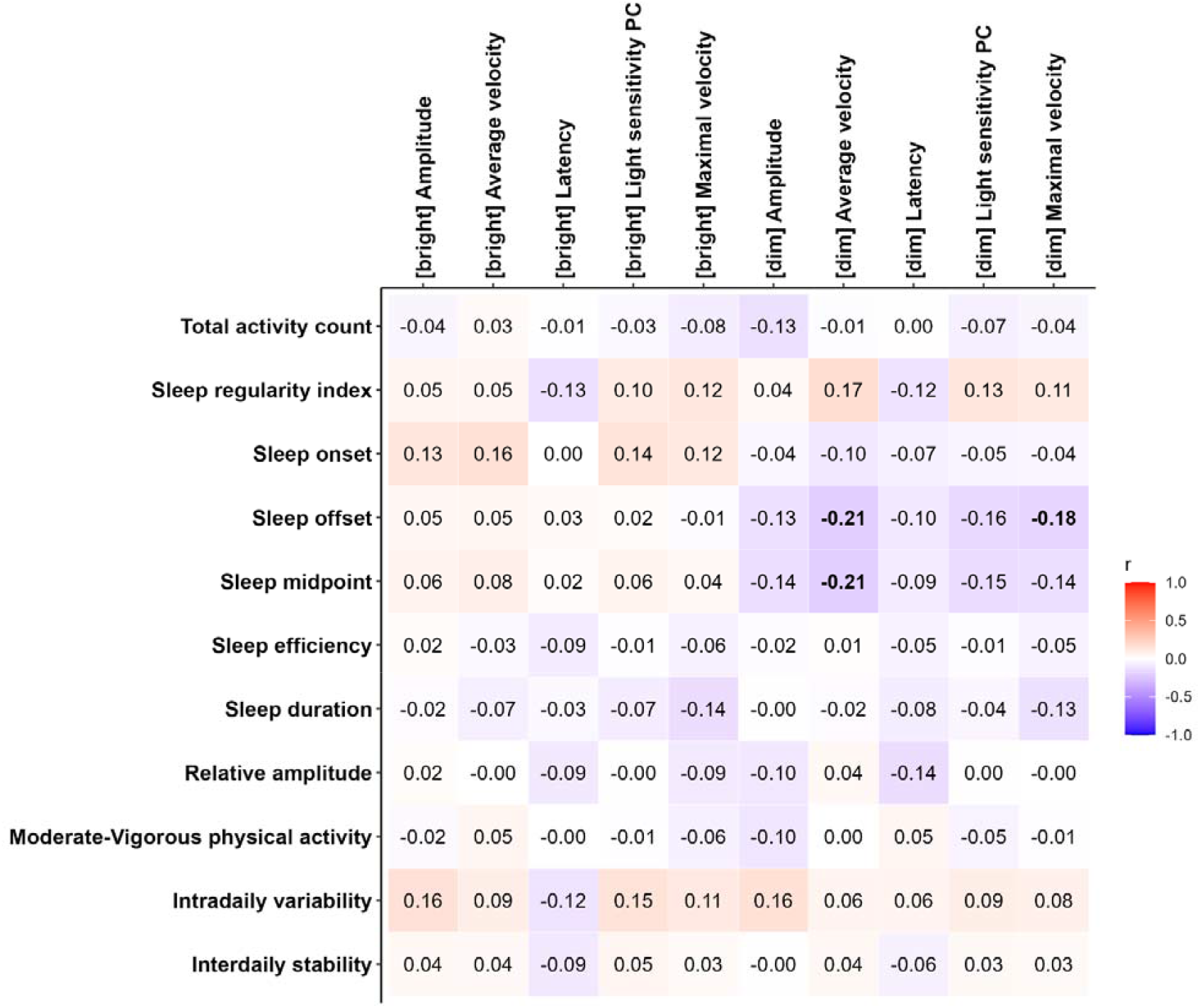
Partial correlation matrix of pupillometry measures and actigraphy-derived measures in the whole sample (N=125). Notes: Values in bold represent significant correlations. Accounting for the effect of age and static pupil diameter.

Maximal constriction velocity after a dim light pulse moderately predicted a subset of young people with delayed sleep offset timing, identified as later than 1 SD from the sample mean, with an area under the ROC curve (AUC) of 0.674 (95% CI = [0.551, 0.798]; sensitivity = 0.684; specificity = 0.642; Youden index threshold = 4.28). Slower maximal constriction velocity after a dim light pulse was associated with delayed sleep offset. When restricting the sample to those not taking mood stabilizers, the association improves modestly (AUC=0.706; 95% CI=[0.577, 0.835]; sensitivity=0.883; specificity=0.5; index threshold = 4.55).

### 3.3 Characteristics of Emerging Mental Disorder Cases

#### 3.3.1 Baseline Assessments

##### 3.3.1.1. Polygenic risk scores

Within cases, bipolar PGS was positively correlated with dim light sensitivity (r=0.28, *p*=0.021), average constriction velocity (r=0.26, *p*=0.033) and maximal constriction velocity (r=0.28, *p*=0.021) after a dim light pulse, indicating that genetic risk for bipolar disorder (BD) was correlated with higher dim light sensitivity. Additionally, Vitamin D PGS was positively correlated with bright light sensitivity (r=0.24, *p*=0.43), average constriction velocity (r=0.24, *p*=0.0.47), and maximal constriction velocity (r=0.26, *p*=0.031). Finally, sleep duration PGS was negatively correlated with amplitude of constriction after a bright light pulse (r=-0.26, *p*=0.033). These results indicates that individuals with higher sensitivity to bright light have higher genetic risk for higher Vitamin D concentration, and lower genetic risk for shorter sleep duration. Details reported in Table S4.

##### 3.3.1.2 Clinical ratings

Within cases, higher insomnia severity (ISI total) was negatively correlated with the average constriction velocity after a dim light pulse (r=-0.17, *p*=0.045) and the latency of constriction after a bright light pulse (r=-0.18, *p*=0.038), indicating lower dim light sensitivity but higher bright light sensitivity was correlated with higher insomnia severity. Higher depressive symptoms (QIDS total) was correlated with decreased latency of constriction after a bright light pulse (r=-0.17, *p*=0.034). Details are reported in Table S5.

##### 3.3.1.3 Medications

Within cases, 26 (17%) were prescribed mood stabilizers and 63 (41%) were prescribed either an SSRI or SNRI. In those prescribed mood stabilizers, the pupil size change after a dim light pulse was smaller (M=1.54 vs M=1.76; F=4.666; *p*=0.032) and latency of constriction was longer (M=0.259 vs M=0.246; F=4.622; *p*=0.033) in cases prescribed mood stabilizers compared to those not prescribed mood stabilizers (but who may have been taking other medications). None of the other pupillometry measures were different. Full comparisons are presented in Figure 3A and Table S6.

**Figure 3.**
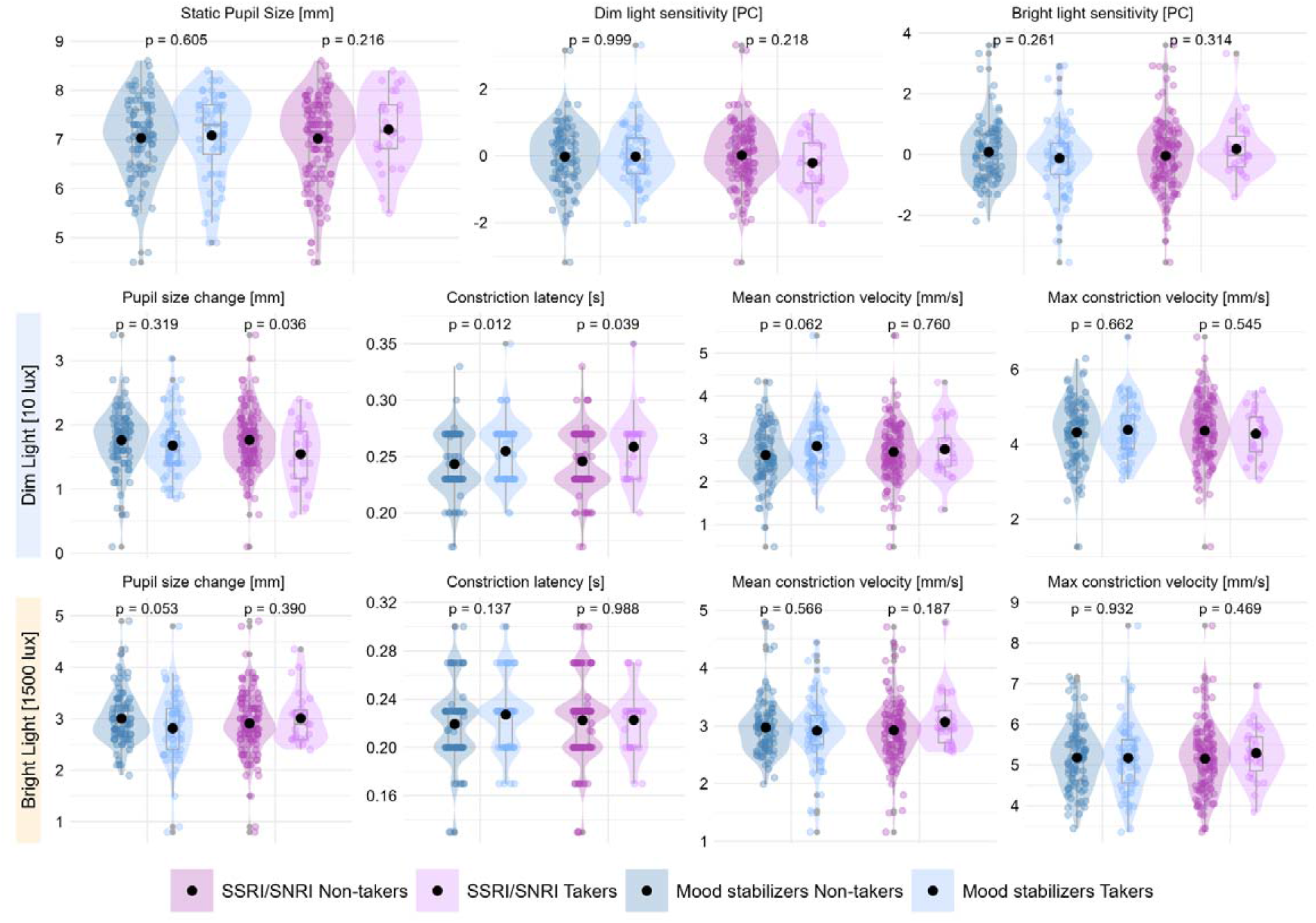
Comparisons of pupillometry measures between clinical cases prescribed (a) mood stabilizers (N=26) vs not prescribed mood stabilizers (N=129); and (b) SSRI or SNRI (N=63) vs. not prescribed SSRI or SNRI (N=92). Note: In each plot, the box represents the interquartile range with median as line inside the box and mean as a black dot with ± 1 standard error as error bars.

In cases prescribed an SSRI or SNRI, The latency of pupil constriction after a dim light pulse was longer (M=0.255 vs M=0.243; F=6.738; *p*=0.010) and amplitude of constriction after a bright light pulse was smaller (M=2.82 vs M=3.01; F=4.012; *p*=0.047) in young people prescribed an SSRI or SNRI compared to those not prescribed either. None of the other pupillometry measures were different. Full comparisons are presented in Figure 3B and Table S7.

#### 3.3.2 Follow-up Assessments of cases

Across all pupillometry measures under dim or bright light, there were no significant changes across visits (which spanned 6-12 month intervals). For overall dim or bright light sensitivity, all visit effects were non-significant (dim: b=-0.02, SE=0.12, t=-0.17, *p=*0.863 for the second visit; b=0.02, SE= 0.23, t=0.10, p=0.918 for the third visit; bright: (b=0.20, SE= 0.12, t=1.71, *p*=0.092 for the second visit, b=-0.09, SE= 0.23, t=-0.37, p=0.710 for the third visit). The same pattern was observed for constriction amplitude (all *p*>0.710), average constriction velocity (all *p*>0.192), maximum constriction velocity (all *p*>0.148) and latency of constriction (all *p>*0.067). Across models, random intercept variances indicated moderate between-person variability (SDs=0.21–0.60). These findings suggest that PLR parameters remained stable across repeated assessments, with no systematic differences between visits in dim or bright light conditions. The stability of PLR measures is represented in Figure 4 and in Table S8.

**Figure 4.**
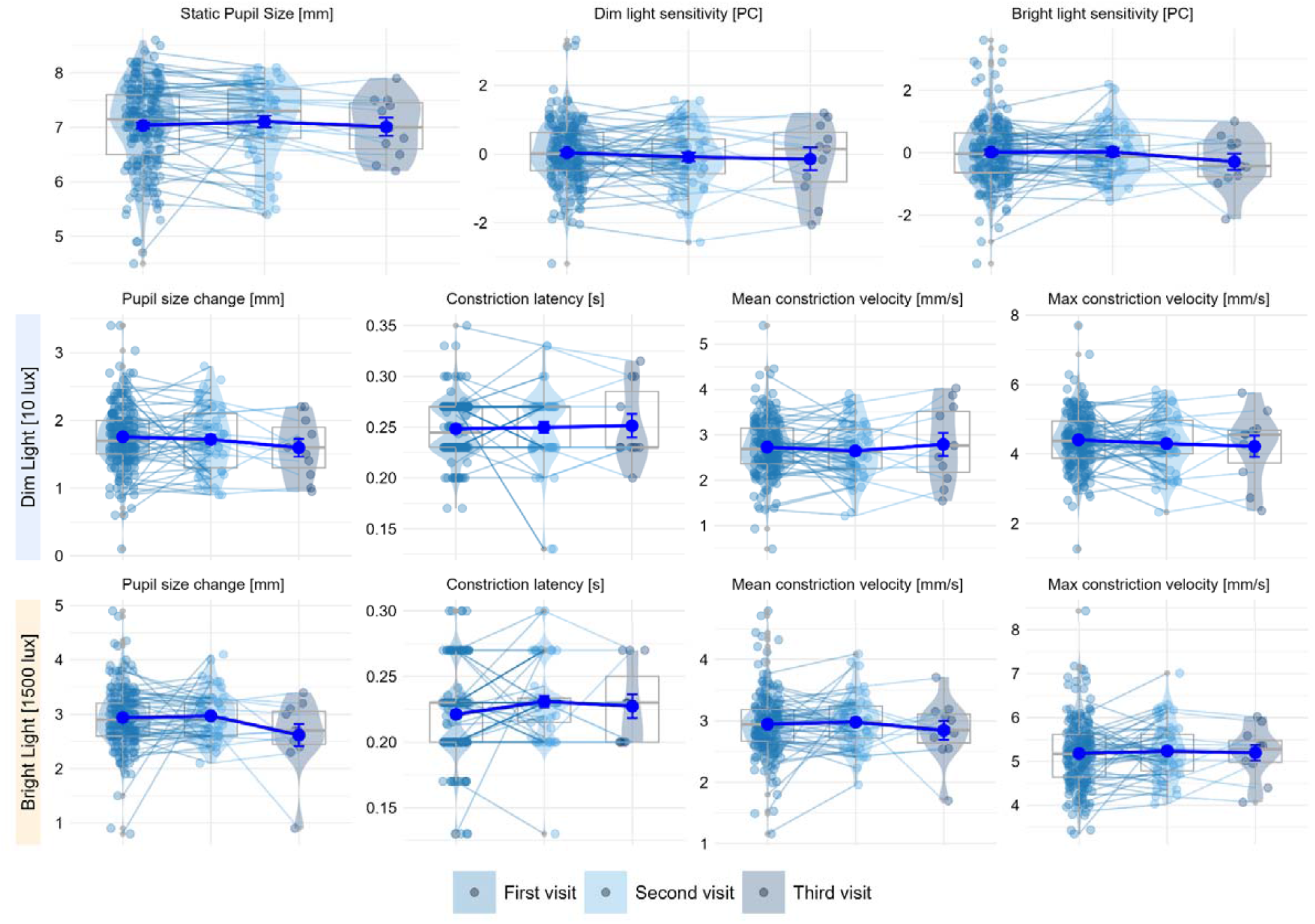
Pupillometry measures across multiple visits (6-12 months apart) (first visit, n=155; second visit, n=49; third visit, n=11). Note: In each plot, the box represents the interquartile range with median as line inside the box and mean as a blue dot with ± 1 standard error as error bars.

## 4 Discussion

Compared to healthy young adults recruited from the same metropolitan community, young people seeking help for mental health at an early intervention service displayed decreased pupil constriction and slower constriction velocity, after a dim light pulse (~10 lux), suggesting lower sensitivity to dim light. In the total sample (i.e., cases and controls), slower pupil constriction velocity after a dim light pulse was correlated with later sleep midpoint and sleep offset times. Further analyses restricted to the cases showed that higher genetic risk for bipolar disorder (BD) was correlated with faster pupil constriction after a dim light pulse, while higher genetic liability for Vitamin D levels was correlated with faster pupil constriction after a bright light pulse. Additionally, while shorter latency of constriction after a bright light pulse was correlated with higher depressive and insomnia symptoms, slower constriction velocity after a dim light pulse was also correlated with higher insomnia symptoms, suggesting lower dim light but higher bright light sensitivity was associated with greater insomnia. Pupillometry measures were stable across time and seasons.

These findings are consistent with studies reporting altered pupillary response in depression, including larger pupil area in darkness and reduced constriction velocity after light pulses in adolescents with major depressive disorder, before exposure to pharmacotherapy (Mestanikova et al., 2017; Wang et al., 2014). Using a different assessment of light sensitivity—melatonin suppression to light—a study reported that circadian light sensitivity was decreased in people with current, but not remitted, depression (McGlashan et al., 2019). Reduced light sensitivity has also been reported in seasonal affective disorder (Roecklein et al., 2013). Interestingly, here, differences between cases and HCs were more pronounced under dim light conditions than bright light, potentially indicating that subtle (i.e., indoor) light exposure may be informative when assessing mood and/or circadian vulnerability. Studies investigating correlates of the PLR are scarce. One study reported that mean constriction velocity after a bright light pulse was the strongest predictor of a delayed-sleep phase disorder phenotype (McGlashan et al., 2018). That study also reported that participants exhibiting circadian disturbances (delayed melatonin timing) had faster PLR responses than both participants with ‘non-circadian’ delayed sleep phase disorder and HCs (McGlashan et al., 2018). Overall, these findings seem to indicate that within some clinical populations, higher sensitivity to light is linked to circadian disturbances, such as delayed sleep phase.

Notably, we also found that higher polygenic risk score (PGS) for BD was correlated with higher sensitivity to dim light. While lower light sensitivity has been associated with unipolar depression (McGlashan et al., 2019; Roecklein et al., 2013), there is converging evidence suggesting that *hyper*sensitivity to light may be a biomarker or risk for relapse in BD (Roguski et al., 2024). Additionally, there is growing interest in investigating retinal integrity in mental health disorders, and BD specifically (Kendall et al., 2025). A recent study reporting that individuals with mental health disorders showed a more pronounced retinal thinning compared to controls (Georgiadis et al., 2025). Additionally, we found a positive correlation between PGS for Vitamin D levels and sensitivity to bright light. Links between higher vitamin D and better mental health and lower insomnia severity have been reported (Singh et al., 2024). Further research is needed to understand the mechanisms linking vitamin D to mental health outcomes.

The effect of mood stabilizers such as lithium on light sensitivity is highly topical. Within our cases, 17% were taking mood stabilizers, and these individuals showed reduced pupil constriction and slower latency of constriction under dim light, although other PLR measures were unaffected. This is consistent with long-standing evidence suggesting that mood stabilizers, and specifically lithium, reduce melatonin light sensitivity (Hallam et al., 2005; Seggie et al., 1989). As a sensitivity analysis, we compared PLR measures between HCs and cases, restricted to those not taking mood stabilizers. Lower maximal constriction velocity in the cases compared to the HCs remained significant, while the constriction amplitude after a dim light pulse was no longer different between groups (Figure S1). Importantly, PLR measures demonstrated high longitudinal stability across multiple visits (spanning 6-12 month intervals) and were not influenced by season (Figure S3, Table S9), indicating that PLR parameters are trait-like and relatively independent of short-term environmental or seasonal variations. This stability underscores the potential utility of PLR measures as reliable biomarkers of individual differences in light sensitivity and circadian physiology.

Perhaps surprisingly, our main finding was that low dim light sensitivity was related to delayed sleep-wake timing. While it has traditionally been thought that increased sensitivity to light leads to delayed timing, emerging evidence seems to suggest that two important factors need to be considered: light exposure and circadian period (Stone et al., 2020). Here, we did not find a mediating effect of light exposure on the relationship between light sensitivity and sleep-wake timing (detailed in Supplementary Materials), which may suggest that other circadian factors play a role or that different composite metrics of light exposure should be investigated. Using a computational model to simulate sleep and circadian timing, a recent study showed that while exposure to nighttime light was particularly impactful in those with high light sensitivity or a long circadian period, bright light in the morning could be maladaptive in those with a short period or low light sensitivity (Stone et al., 2025). Further work is needed to determine how distinct measures influenced by ipRGC function, such as circadian entrainment, PLR, and modulation of mood and alertness modulation, are inter-related. It is likely that some measures have more clinical utility than others, while other might be more influenced by pharmacology than others, highlighting the importance of studying them in tandem.

Several methodological limitations should be mentioned. First, given the exploratory nature of the study, regression analyses were not corrected for multiple comparisons; accordingly, reported associations do not survive correction and require replication. Second, we treated the case sample as a single heterogenous group; larger samples will allow examination of diagnostic or mood disorder subtypes differences. Third, the pupillometry protocol did not allow us to investigate the post-illumination pupil response (PIPR), a melanopsin-driven measure of ipRGC activity (Adhikari et al., 2015; Gamlin et al., 2007). Although alterations in PIPR have been reported in mood disorders, findings are mixed, with evidence for reduced, increased, or unchanged pupil across studies (Berman et al., 2018; Bullock et al., 2019; Chan et al., 2025; Feigl et al., 2018; Laurenzo et al., 2016; Madsen et al., 2021; Roecklein et al., 2013). Although the more acute PLR responses assessed in this study are likely influenced by ipRGC activity, studies investigating light sensitivity in mood disorders should ideally use protocols which allow for the isolation of ipRGC-driven responses (e.g., PIPR) alongside the PLR.

## 5 Conclusion

In conclusion, compared to healthy young adults recruited from the same metropolitan community, young people with emerging mental disorders displayed smaller pupil constriction and slower pupil constriction velocity after a dim light pulse, indicative of a lower sensitivity to dim light. When investigating actigraphy (wearable) correlates of the pupillary light reflex, we found that dim light sensitivity was associated with delayed sleep-wake timing. The prospective repeated-measures design of the study allowed us to demonstrate that pupillometry measures were stable over time and do not appear to be affected by seasons. Using our continuously recruiting *Neurobiology Youth Follow-Up* Study (Nichles et al., 2021), future studies will explore whether PLR measures change alongside clinical states and whether PLR measures predict the future course of illness, and might therefore help differentiate subtypes of mood disorders.

## Data Availability

All data produced in the present study are available upon reasonable request to the authors

## Funding

This work was supported in part by a Mental Health Award from Wellcome Trust (227089_Z_23_Z) and a National Health and Medical Research Council (NHMRC) Synergy Grant (2019260). JJC is supported by an NHMRC Emerging Leadership (EL1) Investigator Grant (GNT2008196). IBH is supported by an NHMRC Leadership (L3) Investigator Grant (GNT2016346). EMM is supported by an NHMRC Emerging Leadership (EL1) Investigator Grant (GNT2033519). This work was also supported by a philanthropic donation from a family affected by mental illness (who would like to be left anonymous).

## Acknowledgements

This work was supported by a National Health and Medical Research Council Synergy Grant (2019260). The study protocol used by 23andMe Research Institute, was approved by an external AAHRPP-accredited institutional review board. We would like to thank the research participants and employees of 23andMe Research Institute, for making this work possible. This work was carried out under SOW7 between 23andMe Research Institute, and the University of Queensland.

## Declarations of interest

IBH is the co-director of Health and Policy at the Brain and Mind Centre at the University of Sydney, which operates early-intervention youth services at Camperdown under contract to Headspace; has previously led community-based and projects supported by the pharmaceutical industry (Wyeth, Eli Lily, Servier, Pfizer, AstraZeneca, Janssen, Cilag), focused on the identification and better management of anxiety and depression; and is the Chief Scientific Advisor to, and a 3·2% equity shareholder in, InnoWell, which aims to transform mental health services through the use of innovative technologies. SWC is Co-Founder of Circadian Health Innovations PTY LTD, a company making wearable light sensors. JWC received travel support from Lundbeck HK. Ltd., and research grants from Takeda. Inc.

## Notes

### Author Declarations

Ethics approval was obtained from the Sydney Local Health District Human Research Ethics Committee (2020/ETH01272) for the Neurobiology study and (X23-0148 & 2023/ETH006188) for control participants. All participants provided written informed consent; for those under 16 years, parental/guardian consent was also obtained.

### Summary of Updates

Typos in authorship list revised.

