## Supplementary Materials for "Dim light sensitivity and delayed sleep timing in young people with emerging mental disorders"

**Table S1 Descriptive statistics of sample with complete actigraphy data**

**Table S2 Partial correlations between pupillometry and actigraphy measures in the whole sample**

**Table S3 Mediation models by light exposure**

**Table S4 Partial correlations between pupillometry measures and PGS score**

**Table S5 Partial correlations between pupillometry measures and clinical scores**

**Table S6 Comparisons of actigraphy data and pupillometry measures between those taking mood stabilizers and those not taking mood stabilizers**

**Table S7 Comparisons of actigraphy data and pupillometry measures between those taking SSRI or SNRI and those not taking either**

**Table S8 Linear mixed effect model of pupillometry measures across multiple visits**

**Table S9 Comparisons of pupillometry measures between seasons**

**Figure S1 Comparisons of pupillometry measures between controls and cases, restricted to those not taking mood stabilizers.**

**Figure S2 Correlation matrix between PLR measures**

**Figure S3 Pupillometry measures by season (Autumn, n=41; Spring, n=42; Summer, n=37; Winter, n=35)**

| **Table S1 Descriptive statistics of sample with complete actigraphy data** | | | | | | |
| --- | --- | --- | --- | --- | --- | --- |
|  |  |  | **Group difference** | | | |
| **Mean** ± SD | **Controls (n=24)** | **Cases (n=101)** | **F/X^2^** | **df** | ***p-value*** | **95% CI** |
| **Age** | 25.67 ± 2.99 | 25.73 ± 5.23 | -0.082 | 61.25 | 0.935 | [-1.67, 1.54] |
| **Sex** [female; N (%)] | 12 (50) | 59 (58) | 0.269 | 1 | 0.604 |  |
| *Pupillometry* |  |  |  |  |  |  |
| **Static Pupil Diameter** [mm] | 6.98 ± 0.6 | 7.04 ± 0.91 | 0.095 | 1,122 | 0.774 | [-0.33, 0.44] |
| *Bright pulse* |  |  |  |  |  |  |
| **Amplitude** [mm] | 2.97 ± 0.47 | 2.9 ± 0.56 | 0.330 | 1,122 | 0.571 | [-0.31, 0.17] |
| **Average CV** [mm/s] | 2.94 ± 0.42 | 2.93 ± 0.5 | 0.012 | 1,122 | 0.918 | [-0.23, 0.20] |
| **Maximal CV** [mm/s] | 5.23 ± 0.63 | 5.12 ± 0.74 | 0.405 | 1,122 | 0.522 | [-0.42, 0.21] |
| **Latency** [s] | 0.21 ± 0.04 | 0.22 ± 0.03 | 1.261 | 1,122 | 0.262 | [-0.01, 0.02] |
| **PC Light Sensitivity** | 0.09 ± 0.86 | -0.05 ± 0.98 | 0.432 | 1,122 | 0.513 | [-0.57, 0.28] |
| *Dim pulse* |  |  |  |  |  |  |
| **Amplitude** [mm] | **1.97 ± 0.48** | **1.75 ± 0.48** | **3.979** | **1,122** | **0.048** | **[-0.43, 0.00]** |
| **Average CV** [mm/s] | 2.84 ± 0.61 | 2.68 ± 0.63 | 1,187 | 1,122 | 0.274 | [-0.43, 0.12] |
| **Maximal CV** [mm/s] | **4.8 ± 0.97** | **4.34 ± 0.76** | **6.382** | **1,122** | **0.013** | **[-0.82, -0.1]** |
| **Latency** [s] | 0.25 ± 0.03 | 0.25 ± 0.03 | 0.042 | 1,122 | 0.837 | [-0.01, 0.01] |
| **PC Light Sensitivity** | 0.39 ± 1.01 | -0.02 ± 0.89 | 3.821 | 1,122 | 0.053 | [-0.82, 0.00] |
| *Note*s:  Abbreviations: CV: constriction velocity; PC: Principal Component-derived; MVPA: Moderate-to-vigorous physical activity. Bold text indicates significant differences. Adjusted for age and uncorrected for multiple comparisons. | | | | | | |

**Table S2. Partial Correlations between pupillometry measures and actigraphy measures in the whole sample**

|  |  |  |  | Dim light | | | | | Bright light | | | | |
| --- | --- | --- | --- | --- | --- | --- | --- | --- | --- | --- | --- | --- | --- |
| *Variable (r)* | N | *M* | *SD* | Amplitude | Latency | ACV | MCV | PC | Amplitude | Latency | ACV | MCV | PC |
| Sleep onset | 125 | 23.65 | 1.59 | -0.04 | -0.07 | -0.10 | -0.04 | -0.05 | 0.13 | 0.00 | 0.16 | 0.12 | 0.14 |
| Sleep offset | 125 | 32.50 | 1.57 | -0.13 | -0.10 | **-0.21*** | **-0.18*** | -0.16 | 0.05 | 0.03 | 0.05 | -0.01 | 0.02 |
| Sleep duration | 125 | 7.11 | 1.11 | -0.00 | -0.08 | -0.02 | -0.13 | -0.04 | -0.02 | -0.03 | -0.07 | -0.14 | -0.07 |
| Sleep efficiency | 125 | 83.62 | 6.16 | -0.02 | -0.05 | 0.01 | -0.05 | -0.01 | 0.02 | -0.09 | -0.03 | -0.06 | -0.01 |
| Sleep midpoint | 125 | 28.20 | 1.48 | -0.14 | -0.09 | **-0.21*** | -0.14 | -0.15 | 0.06 | 0.02 | 0.08 | 0.04 | 0.06 |
| Relative amplitude | 125 | 0.79 | 0.08 | -0.10 | -0.14 | 0.04 | -0.00 | 0.00 | 0.02 | -0.09 | -0.00 | -0.09 | -0.00 |
| Intradaily variability | 125 | 0.77 | 0.15 | 0.16 | 0.06 | 0.06 | 0.08 | 0.09 | 0.16 | -0.12 | 0.09 | 0.11 | 0.15 |
| Interdaily stability | 125 | 0.26 | 0.08 | -0.00 | -0.06 | 0.04 | 0.03 | 0.03 | 0.04 | -0.09 | 0.04 | 0.03 | 0.05 |
| Sleep regularity index | 115 | 65.08 | 14.60 | 0.04 | -0.12 | 0.17 | 0.11 | 0.13 | 0.05 | -0.13 | 0.05 | 0.12 | 0.10 |
| Total activity count | 125 | 37,578.98 | 12,206.74 | -0.13 | 0.00 | -0.01 | -0.04 | -0.07 | -0.04 | -0.01 | 0.03 | -0.08 | -0.03 |
| MVPA | 125 | 83.36 | 47.31 | -0.10 | 0.05 | 0.00 | -0.01 | -0.05 | -0.02 | -0.00 | 0.05 | -0.06 | -0.01 |
| *Note.* Partial correlations controlling for static pupil size and age. **p* < .05. M: mean; SD: standard deviation; AVC: average constriction velocity; MCV; maximal constriction velocity; PC: light sensitivity principal component; MVPA: Moderate to vigorous physical activity. | | | | | | | | | | | | | |

| **Table S3 Mediation models of associations dim light sensitivity and sleep midpoint by light exposure (N=114)** | | | | | | | | |
| --- | --- | --- | --- | --- | --- | --- | --- | --- |
|  |  |  |  | **Predictor** | | | | |
| **IV** | **DV** | **Mediator** | **Path** | **Beta** | **SE** | **Z** | ***p-value*** | **95% CI** |
| Dim ACV | Ligh 3h pre-sleep |  | a | 49.105 | 59.652 | 0.823 | 0.410 | [-88.11, 170.63] |
| Ligh 3h pre-sleep | Sleep midpoint |  | b | **0.000** | **0.000** | **-2.475** | **0.013** | **[-0.001, -0.0001]** |
| Dim ACV | Sleep midpoint |  | c’ (direct) | **-0.501** | **0.231** | **-2.168** | **0.030** | **[-0.921, -0.040]** |
| Dim ACV | Sleep midpoint | Ligh 3h pre-sleep | ab (indirect) | -0.022 | 0.028 | -0.790 | 0.429 | [-0.082, 0.033] |
| Dim ACV | Sleep midpoint |  | c (total) | **-0.523** | **0.237** | **-2.208** | **0.027** | **[-0.956, -0.060]** |
| Dim ACV | Light 3h post-wake |  | a | -41.235 | 224.717 | -0.183 | 0.854 | [-493.95, 327.23] |
| Light 3h post-wake | Sleep midpoint |  | b | 0.000 | 0.000 | 0.177 | 0.859 | [-0.0002, 0,0002] |
| Dim ACV | Sleep midpoint |  | c’ (direct) | **-0.523** | **0.237** | **-2.206** | **0.042** | **[-0.962, -0.010]** |
| Dim ACV | Sleep midpoint | Light 3h post-wake | ab (indirect) | -0.001 | 0.024 | -0.030 | 0.800 | [-0.070, 0.036] |
| Dim ACV | Sleep midpoint |  | c (total) | **-0.523** | **0.237** | **-2.208** | **0.040** | **[-0.960, -0.0500]** |
| Dim ACV | Mean daily light |  | a | 138.515 | 190.005 | 0.729 | 0.466 | [-227.4, 539.3] |
| Mean daily light | Sleep midpoint |  | b | **0.000** | **0.000** | **-4.563** | **0.000** | **[-0.0005, -0.0002]** |
| Dim ACV | Sleep midpoint |  | c’ (direct) | -0.474 | 0.236 | -2.005 | 0.056 | [-0.878, 0.010] |
| Dim ACV | Sleep midpoint | Mean daily light | ab (indirect) | -0.050 | 0.070 | -0.710 | 0.382 | [-0.179, 0.087] |
| Dim ACV | Sleep midpoint |  | c (total) | **-0.523** | **0.237** | **-2.208** | **0.046** | **[-0.950, -0.010]** |
| Notes. In all models, we accounted for the effect of age and the static pupil size. c’ corresponds to the direct effect of IV on DV; c corresponds to the total effect of IV on DV; ab: indirect effect of X on Y via M. ACV: average constriction velocity; CI: confidence interval; SE: standard error. | | | | | | | | |

**Table S4. Partial Correlations between pupillometry measures and polygenic risk score in cases restricted to European ancestry**

|  |  | |  | Dim light | | | | | Bright light | | | | |
| --- | --- | --- | --- | --- | --- | --- | --- | --- | --- | --- | --- | --- | --- |
| *Variable (r)* | N | *M* | *SD* | Amplitude | Latency | ACV | MCV | PC | Amplitude | Latency | ACV | MCV | PC |
| **Bipolar PGS** | 73 | 0.63 | 0.64 | 0.16 | -0.24 | **0.26*** | **0.28*** | **0.28*** | 0.02 | -0.08 | 0.03 | 0.13 | 0.07 |
| **MDD PGS** | 73 | 0.23 | 0.42 | -0.19 | 0.19 | -0.07 | -0.19 | -0.19 | -0.06 | 0.19 | -0.13 | -0.05 | -0.12 |
| **Vitamin D PGS** | 73 | -0.34 | 0.24 | 0.08 | -0.20 | 0.20 | 0.18 | 0.19 | 0.12 | -0.15 | **0.24*** | **0.26*** | **0.24*** |
| **Sleep midpoint PGS** | 73 | 0.10 | 0.11 | -0.09 | 0.04 | 0.05 | -0.03 | -0.03 | -0.03 | 0.17 | -0.03 | 0.08 | -0.02 |
| **Sleep duration PGS** | 73 | -0.11 | 0.17 | -0.06 | 0.07 | -0.21 | -0.05 | -0.12 | **-0.26*** | 0.11 | -0.15 | 0.06 | -0.14 |
| **Chronotype PGS** | 73 | -0.82 | 0.37 | 0.16 | -0.01 | -0.07 | 0.06 | 0.06 | -0.05 | -0.03 | -0.05 | -0.09 | -0.06 |
| **Insomnia PGS** | 73 | 0.02 | 0.10 | -0.03 | -0.06 | 0.04 | -0.06 | -0.01 | 0.21 | 0.02 | 0.19 | 0.13 | 0.18 |
| *Note.* Partial correlations controlling for 2 population ancestry PCs, static pupil size and age. **p* < .05. M: mean; SD: standard deviation; AVC: average constriction velocity; MCV; maximal constriction velocity; PC: light sensitivity principal component; MVPA: Moderate to vigorous physical activity. | | | | | | | | | | | | | |

**Table S5. Partial Correlations between pupillometry measures and clinical scores in cases**

|  |  | |  | Dim light | | | | | Bright light | | | | |
| --- | --- | --- | --- | --- | --- | --- | --- | --- | --- | --- | --- | --- | --- |
| *Variable (r)* | N | M | SD | Amplitude | Latency | ACV | MCV | PC | Amplitude | Latency | ACV | MCV | PC |
| **QIDS Total score** | 155 | 9.25 | 5.23 | 0.01 | -0.09 | 0.01 | 0.07 | 0.05 | 0.01 | **-0.17*** | -0.01 | 0.07 | 0.05 |
| **YMRS Total score** | 155 | 1.01 | 1.71 | -0.02 | -0.08 | 0.05 | -0.01 | 0.02 | -0.00 | 0.01 | -0.01 | -0.00 | -0.01 |
| **OASIS Total score** | 146 | 7.62 | 4.36 | -0.02 | -0.06 | -0.03 | -0.03 | -0.02 | 0.05 | -0.09 | -0.04 | 0.06 | 0.04 |
| **ISI Total score** | 142 | 10.08 | 5.37 | -0.04 | -0.01 | **-0.17*** | -0.10 | -0.11 | 0.03 | **-0.18*** | -0.03 | 0.02 | 0.03 |
| **K10 Total score** | 146 | 26.21 | 7.89 | 0.03 | -0.15 | 0.03 | 0.06 | 0.07 | 0.06 | -0.11 | 0.05 | 0.12 | 0.10 |
| **SOFAS score** | 155 | 69.55 | 12.80 | 0.07 | 0.02 | 0.08 | 0.05 | 0.07 | -0.01 | -0.00 | 0.06 | 0.00 | 0.02 |
| *Note.* Partial correlations controlling for static pupil size and age. **p* < .05. M: mean; SD: standard deviation; AVC: average constriction velocity; MCV; maximal constriction velocity; PC: light sensitivity principal component; MVPA: Moderate to vigorous physical activity. | | | | | | | | | | | | | |

| **Table S6 Comparisons of pupillometry measures between mood stabilizers status** | | | | | | |
| --- | --- | --- | --- | --- | --- | --- |
|  |  |  | **Group differences** | | | |
| **Mean (**SD) | **Non-taker (N=129)** | **Taker (N=26)** | **F** | **df** | ***p-value*** | **95% CI** |
| **Static Pupil Diameter** [mm] | 7.02 (0.83) | 7.20 (0.76) | 1.112 | 1, 152 | 0.290 | [-0.13, 0.57] |
| ***Dim pulse*** |  |  |  |  |  |  |
| **Amplitude** [mm] | **1.76 (0.47)** | **1.54 (0.49)** | **4.666** | **1, 152** | **0.032** | **[-0.42, -0.01]** |
| **Latency** [s] | **0.25 (0.03)** | **0.26 (0.03)** | **4.622** | **1, 152** | **0.033** | **[0.00, 0.02]** |
| **Average CV** [mm/s] | 2.70 (0.66) | 2.76 (0.64) | 0.181 | 1, 152 | 0.670 | [-0.24, 0.32] |
| **Maximal CV** [mm/s] | 4.36 (0.82) | 4.28 (0.64) | 0.246 | 1, 152 | 0.620 | [-0.44, 0.23] |
| **PCA Light Sensitivity** | 0.02 (0.91) | -0.21 (0.80) | 1.390 | 1, 152 | 0.240 | [-0.62, 0.14] |
| ***Bright pulse*** |  |  |  |  |  |  |
| **Amplitude** [mm] | 2.91 (0.60) | 3.01 (0.50) | 0.591 | 1, 152 | 0.440 | [-0.14, 0.36] |
| **Latency** [s] | 0.22 (0.03) | 0.22 (0.03) | 0.002 | 1, 152 | 0.970 | [-0.01, 0.01] |
| **Average CV** [mm/s] | 2.92 (0.54) | 3.07 (0.49) | 1.570 | 1, 152 | 0.210 | [-0.07, 0.38] |
| **Maximal CV** [mm/s] | 5.16 (0.84) | 5.29 (0.67) | 0.595 | 1, 152 | 0.440 | [-0.22, 0.48] |
| **PCA Light Sensitivity** | -0.04 (1.11) | 0.19 (0.93) | 0.926 | 1, 152 | 0.340 | [-0.23, 0.70] |
| *Notes:* Measures are in mm or mm/s. CV: constriction velocity. Accounting for the effect of age. | | | | | | |

| **Table S7 Comparisons of pupillometry measures between SSRI or SNRI intake status** | | | | | | |
| --- | --- | --- | --- | --- | --- | --- |
|  |  |  | **Group differences** | | | |
| **Mean (**SD) | **Non-taker (N=92)** | **Taker (N=63)** | **F** | **df** | ***p-value*** | **95% CI** |
| **Static Pupil Diameter** [mm] | 7.03 (0.80) | 7.08 (0.86) | 0.160 | 1, 152 | 0.690 | [-0.20, 0.33] |
| ***Dim pulse*** |  |  |  |  |  |  |
| **Amplitude** [mm] | 1.76 (0.48) | 1.68 (0.46) | 1.064 | 1, 152 | 0.300 | [-0.23, 0.08] |
| **Latency [s]** | **0.24 (0.03)** | **0.25 (0.03)** | **6.738** | **1, 152** | **0.010** | **[0.00, 0.02]** |
| **Average CV** [mm/s] | 2.62 (0.67) | 2.83 (0.62) | 3.813 | 1, 152 | 0.053 | [-0.01, 0.41] |
| **Maximal CV** [mm/s] | 4.32 (0.85) | 4.39 (0.71) | 0.256 | 1, 152 | 0.610 | [-0.20, 0.31] |
| **PCA Light Sensitivity** | -0.02 (0.91) | -0.02 (0.88) | 0.001 | 1, 152 | 0.970 | [-0.29, 0.29] |
| ***Bright pulse*** |  |  |  |  |  |  |
| **Amplitude** [mm] | **3.01 (0.51)** | **2.82 (0.66)** | **4.012** | **1, 152** | **0.047** | **[-0.37, 0.00]** |
| **Latency** [s] | 0.22 (0.03) | 0.23 (0.03) | 2.282 | 1, 152 | 0.130 | [-0.00, 0.02] |
| **Average CV** [mm/s] | 2.97 (0.49) | 2.91 (0.60) | 0.381 | 1, 152 | 0.540 | [-0.22, 0.12] |
| **Maximal CV** [mm/s] | 5.18 (0.76) | 5.18 (0.89) | 0.003 | 1, 152 | 0.960 | [-0.28, 0.25] |
| **PCA Light Sensitivity** | 0.08 (1.00) | -0.12 (1.20) | 1.328 | 1, 152 | 0.250 | [-0.55, 0.15] |
| *Notes:* Measures are in mm or mm/s. CV: constriction velocity. Accounting for the effect of age. | | | | | | |

| **Table S8 Linear Mixed effect model of pupillometry measures across multiple visits** | | | | | | | |
| --- | --- | --- | --- | --- | --- | --- | --- |
| **Condition** | **Outcome Variable** | **Predictor** | **Estimate (b)** | **SE** | **df** | **t** | **p** |
| **Dim** | PC Light sensitivity | Visit (second) | –0.02 | 0.12 | 98.16 | –0.17 | .863 |
|  |  | Visit (third) | 0.02 | 0.23 | 93.76 | 0.10 | .918 |
|  | Amplitude | Visit (second) | 0.01 | 0.07 | 107.54 | 0.13 | .900 |
|  |  | Visit (third) | –0.004 | 0.13 | 104.47 | –0.03 | .976 |
|  | Average constriction velocity | Visit (second) | –0.05 | 0.08 | 86.70 | –0.58 | .565 |
|  |  | Visit (third) | 0.13 | 0.16 | 81.78 | 0.81 | .420 |
|  | Max constriction velocity | Visit (second) | –0.04 | 0.11 | 111.03 | –0.35 | .724 |
|  |  | Visit (third) | –0.02 | 0.22 | 110.91 | –0.07 | .942 |
|  | Latency | Visit (second) | -0.00 | 0.00 | 129.46 | -0.17 | .892 |
|  |  | Visit (third) | 0.00 | 0.01 | 129.48 | 0.17 | .863 |
| **Bright** | PC Light sensitivity | Visit (second) | 0.20 | 0.12 | 81.78 | 1.71 | .092 |
|  |  | Visit (third) | –0.09 | 0.23 | 77.99 | –0.37 | .710 |
|  | Amplitude | Visit (second) | 0.08 | 0.08 | 139.52 | 1.00 | .321 |
|  |  | Visit (third) | –0.29 | 0.16 | 161.27 | –1.77 | .079 |
|  | Average constriction velocity | Visit (second) | 0.09 | 0.07 | 96.90 | 1.31 | .192 |
|  |  | Visit (third) | –0.03 | 0.14 | 97.22 | –0.22 | .827 |
|  | Max constriction velocity | Visit (second) | 0.13 | 0.09 | 69.46 | 1.46 | .148 |
|  |  | Visit (third) | 0.18 | 0.17 | 64.78 | 1.09 | .280 |
|  | Latency | Visit (second) | 0.01 | 0.00 | 97.58 | 1.84 | .069 |
|  |  | Visit (third) | 0.00 | 0.01 | 98.10 | 0.46 | .650 |
| Notes. Visits were 6 and 12 months apart. | | | | | | | |

| **Table S9 Comparisons of pupillometry measures between seasons** | | | | | | | |
| --- | --- | --- | --- | --- | --- | --- | --- |
|  |  |  |  |  | **Group differences** | | |
| **Mean (SD)** | **Spring**  **(N=50)** | **Summer (N=44)** | **Autumn (N=42)** | **Winter (N=46)** | **F** | **df** | ***p-value*** |
| **Static Pupil Diameter** [mm] | 7.11 (0.76) | 7.04 (0.90) | 6.89 (0.81) | 7.08 (0.70) | 0.660 | 3,177 | 0.580 |
| ***Dim pulse*** |  |  |  |  |  |  |  |
| **Amplitude** [mm] | 1.78 (0.43) | 1.86 (0.51) | 1.75 (0.54) | 1.65 (0.43) | 1.566 | 3, 177 | 0.199 |
| **Latency [s]** | 0.25 (0.03) | 0.25 (0.03) | 0.24 (0.03) | 0.25 (0.03) | 2.219 | 3, 177 | 0.088 |
| **Average CV** [mm/s] | 2.75 (0.77) | 2.73 (0.57) | 2.84 (0.67) | 2.61 (0.53) | 0.896 | 3, 177 | 0.440 |
| **Maximal CV** [mm/s] | 4.45 (0.89) | 4.53 (0.82) | 4.40 (0.76) | 4.25 (0.81) | 0.933 | 3, 177 | 0.426 |
| **PCA Light Sensitivity** | 0.07 (0.97) | 0.13 (0.87) | 0.13 (0.92) | -0.19 (0.87) | 1.331 | 3, 177 | 0.266 |
| ***Bright pulse*** |  |  |  |  |  |  |  |
| **Amplitude** [mm] | 2.88 (0.57) | 2.84 (0.57) | 3.06 (0.57) | 2.98 (0.55) | 1.277 | 3, 177 | 0.284 |
| **Latency** [s] | 0.23 (0.03) | 0.22 (0.03) | 0.22 (0.03) | 0.22 (0.04) | 0.808 | 3, 177 | 0.491 |
| **Average CV** [mm/s] | 2.90 (0.52) | 2.93 (0.57) | 3.07 (0.50) | 2.91 (0.48) | 1.023 | 3, 177 | 0.384 |
| **Maximal CV** [mm/s] | 5.18 (0.80) | 5.12 (0.71) | 5.38 (0.91) | 5.07 (0.71) | 1.236 | 3, 177 | 0.298 |
| **PCA Light Sensitivity** | -0.09 (1.00) | -0.06 (1.08) | 0.28 (1.09) | -0.05 (1.02) | 1.172 | 3, 177 | 0.322 |
| Notes. CV: Constriction velocity. Accounting for the effect of age | | | | | | | |

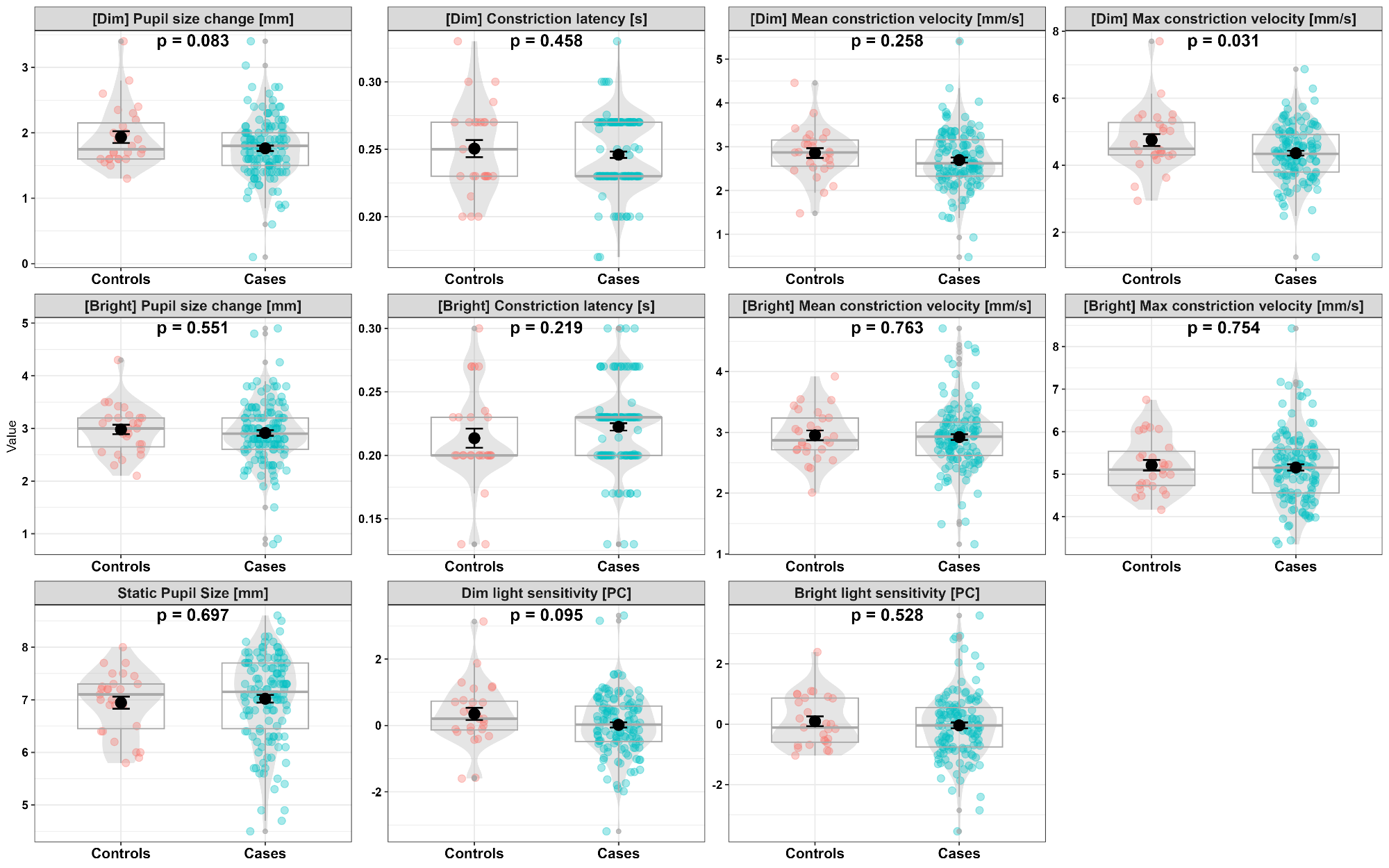
**Figure 3. Comparisons of pupillometry measures between clinical cases prescribed not prescribed mood stabilizers (N=129); and controls (N=27).** Note: In each plot, the box represents the interquartile range with median as line inside the box and mean as a black dot with ± 1 standard error as error bars

**Figure S2 Correlation matrix of PLR measures.** Notes: ACV: average constriction velocity, MCV: maximal constriction velocity. Time interval corresponds to the interval of time between participant’s sleep midpoint and time of pupillometry assessment.

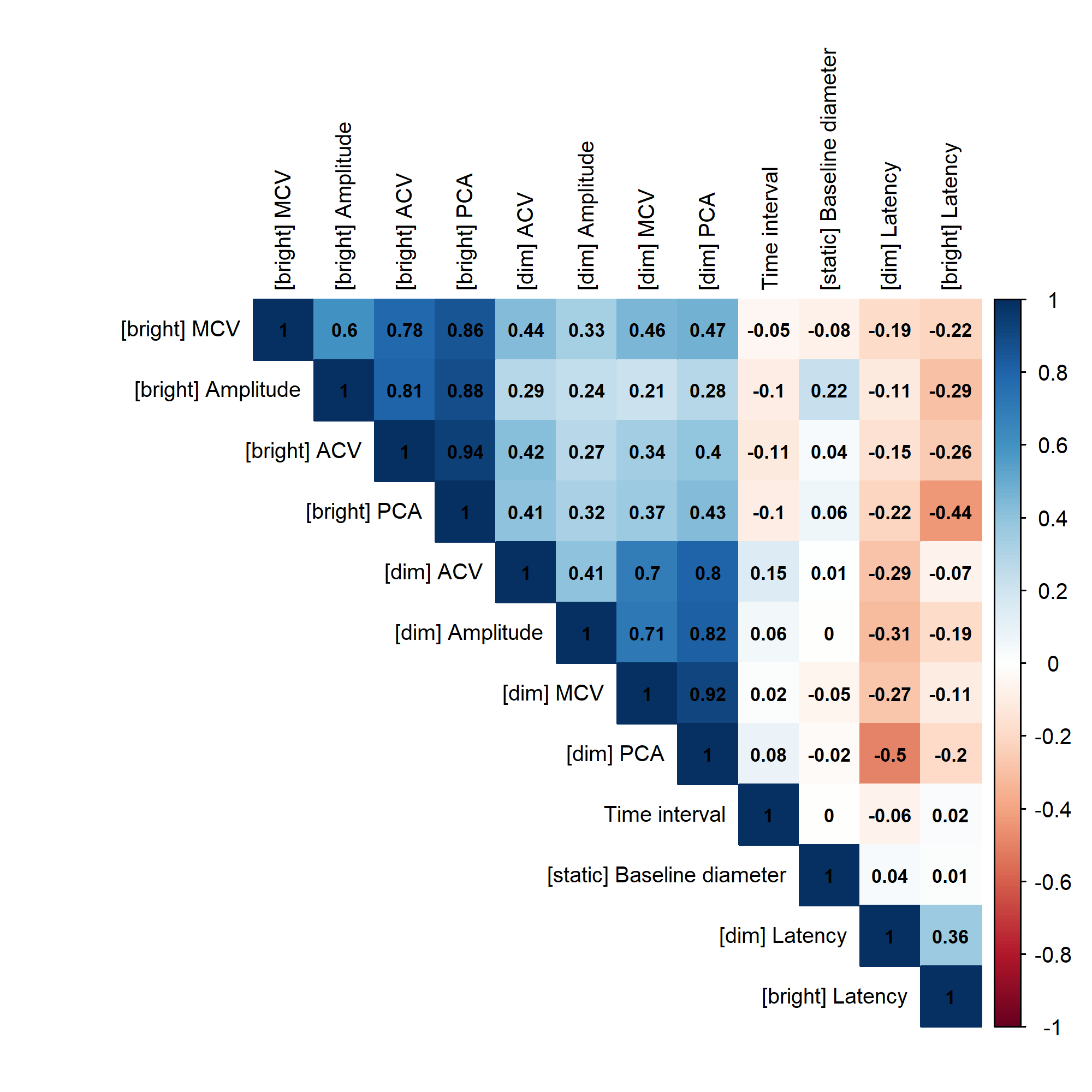

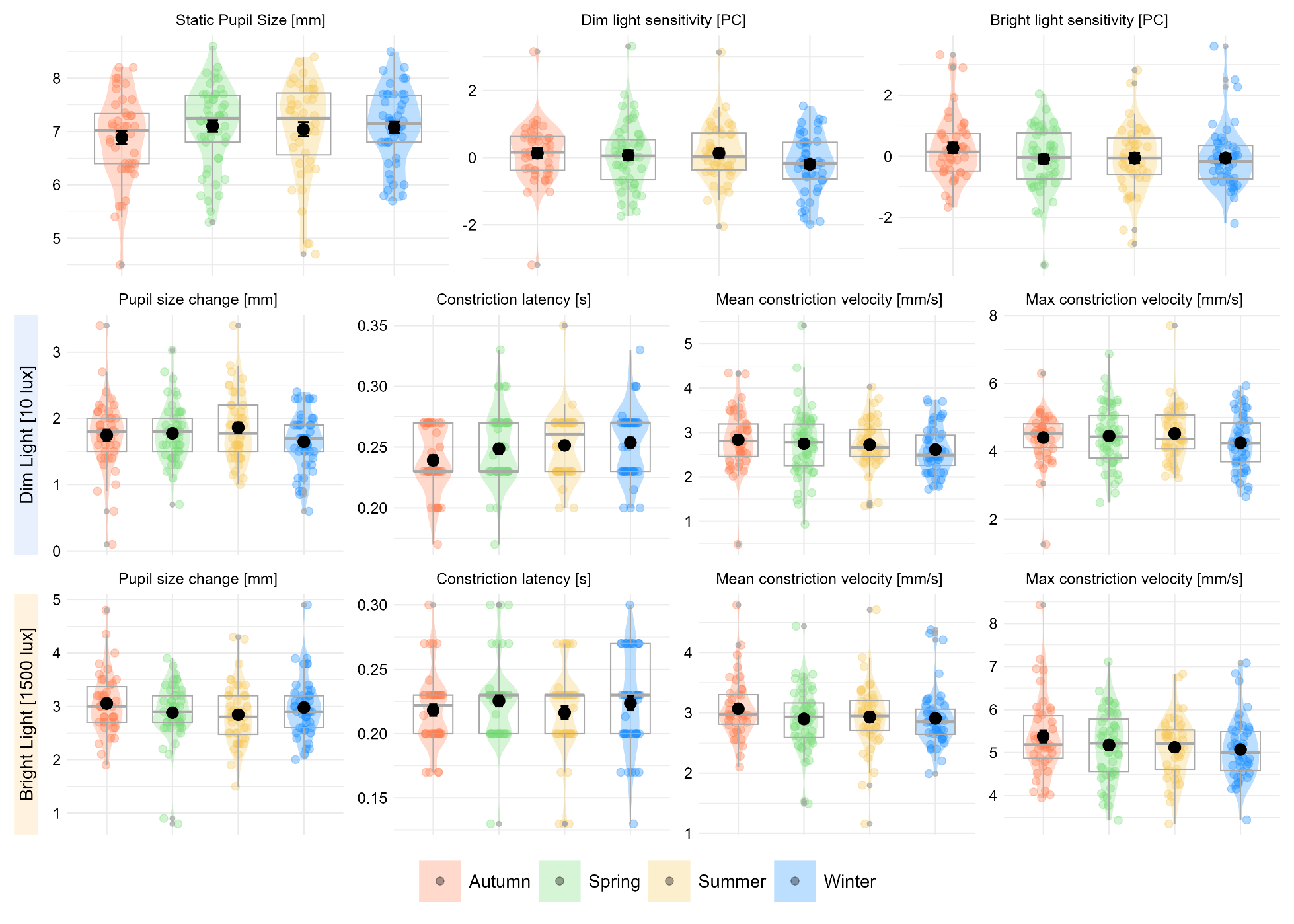
**Figure S3. Pupillometry measures by season (Autumn, n=41; Spring, n=42; Summer, n=37; Winter, n=35).** Note: In each plot, the box represents the interquartile range with median as line inside the box and mean as a black dot with ± 1 standard error as error bars.

**Light exposure metrics**

Light exposure data was obtained from the actigraphy recording device (GENEActiv). Peak light exposure intensity in lux was sampled in 1 minute epoch from the GGIR raw output. Further quality controls step were conducted including (1) discarding invalid epochs (determined by the device), (2) discarding valid epochs with light intensity less than 1 when participant is awake, (3) removing entire days with more than 80% of all epochs with light intensity lower than 10 lux, (4) removing entire days with more than 75% of all epochs with light intensity above 6000 lux, (6) removing entire days with more than 80% of all epochs with the same light intensity value. Additionally, only participants with valid data for at least one weekend day and two weekdays were included.

Following quality controls steps, three metrics were calculated per participant and used in the analyses: average light exposure during the 3h before sleep onset, average light exposure during the 3h after sleep offset (wake time), and average daily light exposure (between sleep offset and sleep onset). Light metrics were calculated per day and then averaged per participant.

**Symptoms scales**

In young people with mood disorders, the following symptoms scales were measured:

- Overall Anxiety Severity and Impairment Scale (OASIS): a 5-item brief dimensional measure of overall anxiety symptoms (Campbell-Sills et al., 2009). The total score was used (range 0-20).
- Quick Inventory of Depressive Symptomatology (QIDS) (Rush et al., 2003): a clinician-administered 16-item inventory assessing the presence of major symptoms of depression, including sleep disturbances, sad mood, change in appetite and weight, poor concentration, suicidal thoughts, lower interest and energy, over the past seven days. Symptoms are rated on a 4-point Likert scale, and a total score ranging from 0 to 27 is derived. A score above 16 indicates severe depression. The total score was used.
- Social and Occupational Functioning Assessment Scale (SOFAS) (Goldman et al., 1992): an observer-rated scale providing a global rating of current functioning with a good construct validity, inter-rater reliability and predictive validity. It ranges from 1 (“*Unable to function without external supervision*”) to 100 (“*Superior functioning*”). The total score was used.
- Young Mania Rating Scale (YMRS) (Young et al., 1978): an observer 11-item assessment measuring the severity of hypo/manic symptoms. Severity of each symptom is rated on a five-point Likert scale, ranging from 0 to 60. The total score was used.
- Insomnia Severity Index (ISI) (Morin et al., 2011): a 7-item self-report questionnaire measuring insomnia. The total score was used, (range 0-7).
- Kessler Psychological Distress Scale (K10) (Kessler et al., 2003): a 10-item self-report scale measuring psychological distress. The total score was used (range 0-10).

**Results: Mediation by light exposure**

To further explore the relationships between slower average constriction velocity after a dim light pulse and later sleep midpoint and sleep offset, we investigated the role of three light exposure metrics (light exposure 3h pre-sleep, light exposure 3h post-wake, and average daily light exposure) as mediators. The direct effect of average constriction velocity after a dim light pulse on sleep midpoint and on sleep offset was significant in all models (all *p<*0.048), but none of the indirect effects via light exposure metrics were significant, indicating that light sensitivity measures appear to relate directly to sleep timing, and not indirectly through light exposure. Light descriptives are reported in Table 1.

**Results: Seasonal variation**

Finally, there were no significant effects of season of assessment on any PLR measures under the dim or bright light conditions, including overall light sensitivity, constriction amplitude, average constriction velocity, or maximum constriction velocity, or latency of constriction (all *p*>0.18). These results suggest that seasonal changes in ambient light exposure did not significantly influence the PLR in this clinical cohort. Full comparisons are presented in Figure S3 and in Table S9.
